# Assessment of Gaming Practices and its effect on scholastic performance of medical students in India: A Case Control study

**DOI:** 10.1101/2020.07.20.20156786

**Authors:** M S Deodatt, R Divya, J Anju, V Raghuram, R Goyal

**Author notes:** The corresponding author’s name, mailing address, and email address. The corresponding author should be aware of the fact that his or her email address can be published or state that no such permission will be granted. Name: Dr Deodatt M Suryawanshi, Mailing Address: Department of Community Medicine, Trichy SRM Medical College Hospital and Research Centre, SRM Nagar, Trichy, Tamil Nadu, India – 621105, Email id /. Source(s) of funding support (e.g., grant funding).: Nil.

## Abstract

**Introduction:** Gaming is a billion-dollar industry growing at a Compound annual growth rate (CAGR) of 9 %-14.3% with biggest market in South East Asian countries. Availability of Low-cost smart phones, ease of internet access has made gaming popular among youth who enjoy it as a leisure activity. According to the WHO excessive indulgence in Gaming can lead to Gaming disorder. Medical students indulging in excessive gaming can succumb to gaming disorder which can affect their scholastic performance. Hence this study was done to assess the gaming practices and its effect on scholastic performance.

**Objective:**

1. To assess the various Gaming practices and the Prevalence of Gaming addiction among medical students.
2. To study the effect of Gaming practices on Scholastic performance of medical students.

**Methods:** The present study used a case control design where the 448(N) study participants were recruited using non probability sampling technique. 91 (Nc) cases who were Gaming for past 6 months were identified using rapid preliminary survey .91 controls (Nco) who never played games were selected and matched for age and sex. Internal Assessment scores (%) of cases and controls were compared. Snedecor F test and Student t test were used to find out the association between the hours of gaming and internal assessment scores (%) and difference of Internal assessment scores between cases and controls respectively. Odds ratio was calculated to identify the risk of Poor scholastic performance. Prevalence of Gaming addiction was assessed using Lemmen’s Gaming addiction scale (GAS).

**Results:** Frequency of gaming (hrs) was not associated with the Mean internal assessment scores (p>0.05). Male students (cases) showed significant reduction in both their internal assessment scores (p<0.001,<0.01) whereas no reduction was observed in Female cases. A negative correlation was observed between GAS and internal assessment scores (r=-0.02). Prevalence of Gaming addiction using GAS was found to be 6.2% among the study population(N=448) and 31% among Cases (Nc=91). The risk of low scores was (*OR =*1.80-1.89) times more in cases than controls.

**Conclusions:** Excessive Gaming adversely affects scholastic performance in males than females. Awareness about Gaming addiction needs to be created among students, parents and teachers. Institutionalised De -addiction services should be made available to medical students.

## Introduction

In 2019 the global gaming market was valued at USD 151.55 billion growing at a compound Annual Growth Rate (CAGR) of 9.17% and expected to reach USD 256.97 billion by 2025 with the largest market in Asia Pacific Smartphone(mobile) remained the most used Gaming platform earning highest USD 64.4 Billion in 2019 ^1^. India is also a rapidly growing Gaming market with an annual growth rate of 14.3 % valued at USD 890 million currently^2^. The growth is driven by rising younger population, higher disposable incomes, introduction of new gaming genres, and the increasing number of smartphone and tablet users^2^.

Though being a harmless leisure activity excessive indulgence in gaming can lead to possible Internet Gaming addictions ^3^.

In the 11th Revision of the International Classification of Diseases (ICD-11) WHO has recognised excessive gaming as a disorder “characterized by impaired control over gaming, increasing priority given to gaming over other activities to the extent that gaming takes precedence over other interests and daily activities, and continuation or escalation of gaming despite the occurrence of negative consequences”4

Studies have documented significant impairment of physical, psychological, social, and work-related problems such as insomnia, increased irritability and aggression, depressive and/or anxiety symptoms, poor academic performance, and neglect of interpersonal relationships with excessive and problematic gaming ^5,6,7^.

It has been found that addiction tendencies are consistently negatively related to scholastic performance. Medical curriculum is vast and requires extensive reading. Gaming in medical students is unexplored so far. This study area would shed light on their Gaming practices and how it affects their scholastic performance.

Hence this study was conducted with the following objectives

1. To study the various Gaming practices among medical students.
2. To assess the prevalence of Gaming disorder among medical students.
3. 3.To study the effect of Gaming practices on Scholastic performance of medical students.

## Material and Methods

The present study made an attempt to demonstrate the of Effect of Gaming on the Scholastic Performance of medical students.

### Study Design

The present study was a Case Control study conducted during the period of October and November 2019 in tertiary Medical college in Trichy district of Tamil nadu,India.Three batches of 448 undergraduate medical students from first year to pre-final year were included as study participants using non probability sampling. Those medical students who were found Gaming (mobile/ pc / internet) for the past 6 months were included as cases in the present study using a rapid preliminary survey. Following the preliminary survey, 91(Nc) students who replied positively for playing games in the last 6 months were taken as cases and 91 (Nco) students who never played games for last 6 months were taken controls. The controls selected were matched for age and sex and the cases were assessed for frequency of gaming hours per week. Internal Assessment scores (%) of last two Internals examinations were accessed from the medical education department of the institution after obtaining written permission from the students. The Internal assessment scores (%) obtained were recorded in percentage and were compared between cases and control to find out significant difference in scores of Cases and Controls.

To assess the prevalence of gaming disorder, Lemmens Gaming addiction scale (GAS) was employed. The Lemmens Gaming addiction scale (GAS) is a pre-tested, pre validated scale with a Cronbach alpha of 0.82 to 0.87 ^8^. It has 7 items such as salience, tolerance, mood modification, relapse, withdrawal, conflict and problems. Each Items has three questions with score range of 0 to 5 with all the components scored on a likert scale, never with score 1, rarely with 2, sometimes with 3, often with 4 and very often with 5. The investigators used the monothetic format in the present study i.e. score of above 3 in all items is indicative of Gaming addiction. Lemmen has himself hypothesized that the monothetic format would lead to a better estimate of the prevalence of addiction than the polythetic format would ^8^. For convenience, the investigators summed up the total score of all 7 items and those having a score of 63 or above was classified to have Gaming addiction.

### Ethical issues

Ethical principles such as respect for the person, and confidentiality were strictly adhered. Institutional Ethics clearance was obtained from Trichy SRM Medical College (IEC Code No: 1007/TSRMMCH&RC/ME-1/2019-IEC no :039)). Informed written consent was obtained from all the participants. If they were absent, they were followed up according to their convenience. The purpose of the study was explained in detail and assured that the results would be used only for scientific purpose.

### Statistical Analysis

The data entry and analysis were done using SPSS software V 21. Descriptive statistics were used for analysing socio-demographic details, frequency and type of gaming. Snedecor F test and Student t test were used to find out the association between the hours of gaming and scholastic performance and Gaming with their internal assessment scores respectively.(N)

## Results

In the present study, out of 448 students who were preliminarily surveyed, 91 were cases (Nc) and 91(Nco) controls. Out of the 91 cases, 49 (53.8%) were female and 42(46.2%) were male. In the present study 70(76.9%) of the cases were below twenty years. Out of 91 cases, 87(94.5%) used mobile phone to play games, 3(4.4%) used a personal computer or laptop and 1(1.1%) used X-Box. (Table.1).

The frequency of playing games was assessed for a typical working day in hours and then calculated for a week. In the present study, 32(35.2%) of Cases played games for more than 8 hours per day, 24(26.4%) for less than 2hrs, 18(19.8%) played for 5 -7 hrs and 17(18.7%) played for 3-4 hrs. (Table 1)

**Table 1:** Age distribution and Gaming Characteristics of Cases. (Nc=91)

|  |  | **Female**<br>**(n, %)** | **Male**<br>**(n, %)** | **Total** | **P value** |
| --- | --- | --- | --- | --- | --- |
| **Age**<br>**distribution**<br>**(yrs.)** | <= 18.0 | 7 (7.6) | 3(3.3) | 10 |  |
|  | 19.0 - 23.0 | 42 (46) * | 38 (41.7) | 80 |  |
|  | 24.0+ | 0 (0) | 1 (1.1) | 1 |  |
| **Platform used**<br>**for Gaming** | Mobile | 48 (52.7)<br>* | 39 (42.8) | 87 | P > 0.05 |
|  | Mobile/Pc | 1(1.1) | 2 (2.1) | 3 |  |
|  | Xbox | 0 (0) | 1 (1.1) | 1 |  |
| **Hours /week**<br>**indulged in**<br>**Gaming (hrs)** | <= 10.0 | 12 (13.1) | 16 (17.5) | 28 | P > 0.05 |
|  | 10.1 - 25.0 | 28(30.7) * | 22(24.1) | 50 |  |
|  | 25.1 - 40.0 | 5(5.4) | 2(2.1) | 7 |  |
|  | 40.1 - 55.0 | 3(3.2) | 0(0) | 3 |  |
|  | 55.1+ | 1(1.1) | 2 (2.1) | 3 |  |
| **Gaming**<br>**Addiction**<br>**Score (GAS)** | <= 62.0 | 34 (37.3)<br>* | 29(31.8) | 63 | P > 0,05 |
|  | 63.0+ | 15(16.4) | 13(14.2) | 28 |  |

\* Highest in the group

There was no significant difference observed in the internal assessment scores of those who played games for more hours than those who played for less hours. (p>0.05) (Table 2).

**Table 2.** Association of Gaming hours with scholastic performance (Nc = 91)

| Hours of gaming/day | Frequency (%) | Mean internal assessment scores | p value* |
| --- | --- | --- | --- |
| **<= 2hrs** | 24(26.4) | 45.7 | **>0.05** |
| **3 to 4hrs** | 17(18.7) | 56.8 |  |
| **5 to 7 hrs** | 18(19.8) | 48.9 |  |
| **>= 8hrs** | 32(35.2) | 49.3 |  |

\*Snedecor F test was used to derive p-value.

In the present study, Male cases(Mc) showed significant reduction in both their internal assessment scores as compared to Male Controls (MCo) (1^st^ Internal assessment p<0.001) and (2nd internal assessment p<0.01). Scholastic scores were unaffected in Female cases (Fc) and controls (Fco) in both the internal assessments s (p>0.05). (Table 3)

**Table 3.**
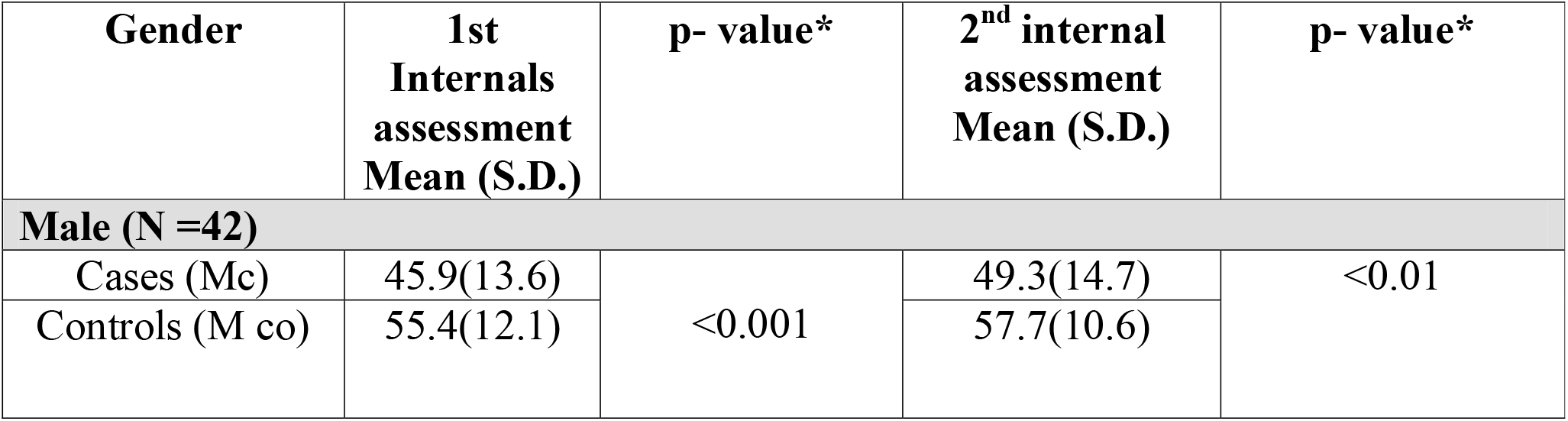
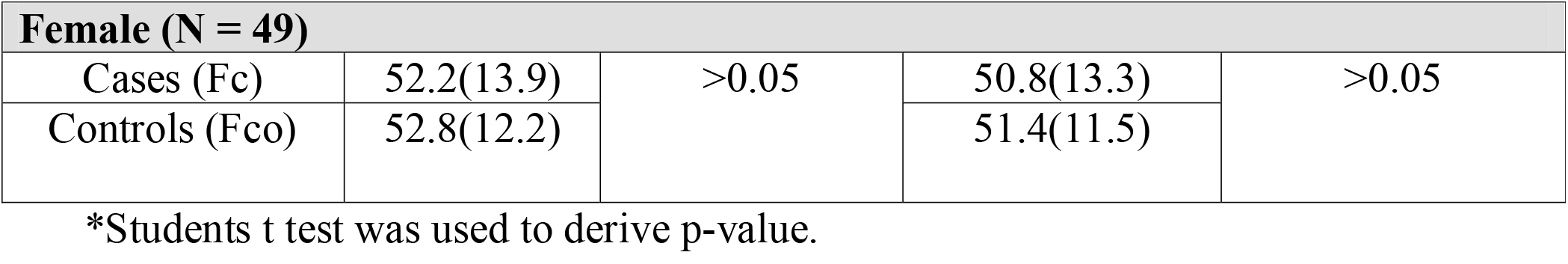
Association of Gender with Internal Assessment marks (Nc= 91)

| Gender | 1st Internals assessment Mean (S.D.) | p- value* | 2<sup>nd</sup> internal assessment Mean (S.D.) | p- value* |
| --- | --- | --- | --- | --- |
| **Male (N =42)** |  |  |  |  |
| Cases (Mc) | 45.9(13.6) | <0.001 | 49.3(14.7) | <0.01 |
| Controls (M co) | 55.4(12.1) |  | 57.7(10.6) |  |

| **Female (N = 49)** |  |  |  |  |
| --- | --- | --- | --- | --- |
| Cases (Fc) | 52.2(13.9) | >0.05 | 50.8(13.3) | >0.05 |
| Controls (Fco) | 52.8(12.2) |  | 51.4(11.5) |  |

\*Students t test was used to derive p-value.

The 7 Items of Lemmens Gaming addiction scale (GAS) were analysed in cases (Nc=91). Salience score > = 3 was observed in 30(33%),tolerance score of >=3 in 24(26.4%), mood modification score >=3 in 34(37.4%), relapse score >=3 in 20 (22%),withdrawal score > = 3 in 26(28.6%) conflict scores of >=3 in 22(24.2%) and Problem scores of > = 3 in 56(61.5%) in cases.(Table 4).

**Table 4.** Scoring of Items in the Gaming Addiction Scale (Nc= 91)

|  | How often during the last six months... | Never | Rarely | Sometimes | Often | Very often |
| --- | --- | --- | --- | --- | --- | --- |
|  | Items | 1 (%) | 2 | 3 | 4 | 5 |
| **A** | **Salience** |  |  |  |  |  |
| **1** | Did you think about playing a game all day long? | 40<br>( 44%) | 16<br>(17.6%) | 16<br>( 17.6%) | 13<br>(14.3%) | 6<br>(6.6%) |
| **2** | Did you spend much free time on games? | 17<br>(18.7%) | 23<br>(25.3 %) | 20<br>( 22%) | 21<br>(23.1%) | 10<br>(11%) |
| **3** | Have you felt addicted to a game? | 18<br>(19.8%) | 18<br>(19.8 %) | 29<br>(31.9%) | 16<br>(17.6%) | 10<br>(11%) |
| **B** | **Tolerance** |  |  |  |  |  |
| **1** | Did you spend increasing amounts | 25 | 16 | 28 | 16 | 6 |
|  | of time on games? | (27.5%) | (17.6%) | (30.8%) | (17.6%) | (6.6%) |
| 2 | Did you play longer than intended? | 20<br>(22%) | 16<br>(17.6%) | 22<br>(24.2%) | 25<br>(27.5%) | 8<br>(8.8%) |
| 3 | Were you unable to stop once you started playing? | 29<br>(31.9) | 15<br>(16.5%) | 25<br>(27.5%) | 13<br>(14.3%) | 9<br>(9.9%) |
| **C** | **Mood Modification** |  |  |  |  |  |
| 1 | Did you play games to forget about real life? | 31 (34.1%) | 15<br>(16.5%) | 20<br>(22%) | 18<br>(19.8%) | 7<br>(7.7%) |
| 2 | Have you played games to release anger or stress? | 26 (28.6%) | 11<br>(12.1%) | 25<br>(27.5%) | 20 (22%) | 9 (9.9%) |
| 3 | Have you played games to feel better? | 20 (22%) | 12<br>(13.2%) | 24<br>(26.4%) | 25<br>(27.5%) | 10 (11%) |
| **D** | **Relapse** |  |  |  |  |  |
| 1 | Have others unsuccessfully tried to reduce your game use? | 29 (31.9%) | 24<br>(26.4%) | 16<br>(17.6%) | 14<br>(15.4%) | 8<br>(8.8%) |
| 2 | Were you unable to reduce your game time? | 33 (36.3%) | 24<br>(26.4%) | 21<br>(23.1%) | 10 (11%) | 3 (3.3%) |
| 3 | Have you failed when trying to reduce game time? | 34 (37.4%) | 20 (22%) | 17<br>(18.7%) | 14<br>(15.4%) | 6 (6.6%) |
| **E** | **Withdrawal** |  |  |  |  |  |
| 1 | Have you felt bad when you were unable to play? | 35 (38.5%) | 22<br>(24.2%) | 13<br>(14.3%) | 16<br>(17.6%) | 5 (5.5%) |
| 2 | Have you become angry when unable to play? | 44 (48.4%) | 19<br>(20.9%) | 14<br>(15.4%) | 9 (9.9%) | 5 (5.5%) |
| 3 | Have you become stressed when unable to play? | 47 (51.6%) | 16<br>(17.6%) | 15<br>(16.5%) | 10 (11%) | 3<br>(3.3) |
| **F** | **Conflict** |  |  |  |  |  |
| 1 | Did you have fights with others (e.g., family, friends) over your time spent on games? | 42 (46.2%) | 21<br>(23.1%) | 15<br>(16.5%) | 10 (11%) | 3 (3.3%) |
| 2 | Have you neglected others (e.g., family, friends) because you were playing games? | 38 (41.8%) | 23<br>(25.3%) | 18<br>(19.8%) | 9 (9.9%) | 3 (3.3%) |
| **3** | Have you lied about time spent on games? | 35 (38.5%) | 21 (23.1) | 21 (23.1%) | 10 (11%) | 4 (4.4%) |
| **G** | **Problems** |  |  |  |  |  |
| **1** | Have you neglected other important activities (e.g., school, work, sports) to play games? | 38 (41.8%) | 19 (20.9) | 17 (18.7%) | 12 (13.2%) | 5 (5.5%) |
| **2** | ...has your time on games caused sleep deprivation? | 34 (37.4%) | 17 (18.7%) | 23 (25.3%) | 9 (9.9%) | 8 (8.8%) |
| **3** | ...did you feel bad after playing for a long time? | 33 (36.3%) | 13 (14.3%) | 24 (26.4%) | 14 (15.4%) | 7 (7.7%) |

In the present study, out of the 448(N) total students who were surveyed, 28 cases (Nc) had a Gaming addiction score (GAS) of = > 63. Thus, the prevalence of Gaming addiction in the present study was observed to be 6.2% (N=448) and 31% among cases (Nc=91).

There was a significant difference observed between mean GAS scores of Male case and Female cases (Table 1, Table 5). GAS scores of Female cases were 9 percentage points more than males’ cases.

**Table 5.** Gender wise comparison of Gaming Addiction score.

|  | **N** | **Mean GAS scores** | **SD** | **p value** |
| --- | --- | --- | --- | --- |
| **Male(Mc)** | 13 | 69.5 | 6.4 | 0.008 |
| **Female(Fc)** | 15 | 78.5 | 9.2 |  |

There was a negative correlation observed between GAS and Mean internal assessment scores for the cases. (r= - 0.02).

**Fig 1.**
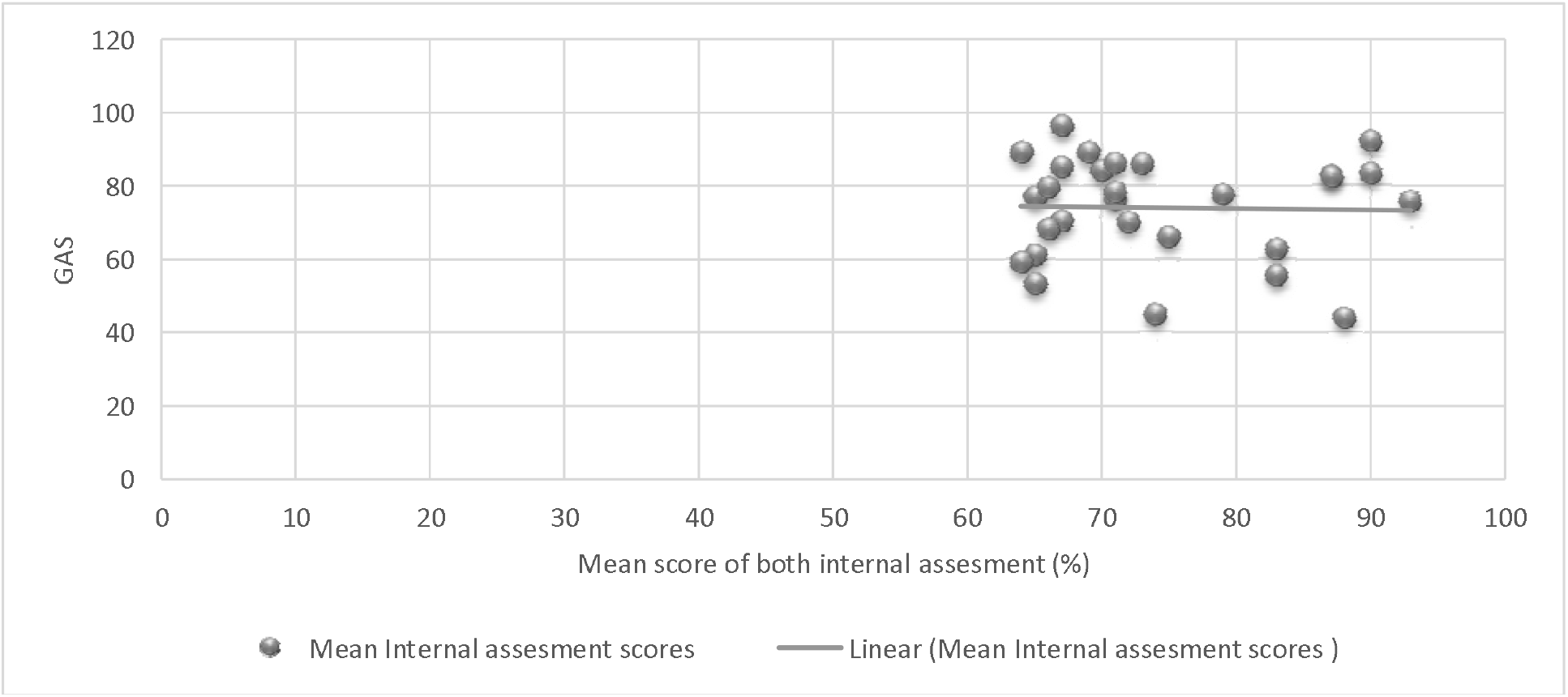
Relationship between Mean internal assessment scores and GAS (r= −0.02).

In the present study it was observed that the odds of students scoring less than 50% was 1.89 times more among cases than controls. This was observed even during the 2^nd^ Internal assessment. (Table 6)

**Table 6.** Risk estimation in Cases and Controls Using Odds ratio

|  | Internal<br>Assessment scores<br>(%) | Cases | Controls | Total | OR |
| --- | --- | --- | --- | --- | --- |
| 1st internal<br>Assessment | < 50 | 45 | 31 | 76 | 1.89 |
|  | > 50 | 46 | 60 | 106 |  |
|  | Total | 91 | 91 | 182 |  |
| 2nd<br>internal<br>Assessment | Internal<br>Assessment scores<br>(%) | Cases | Controls | Total | OR |
|  | < 50 | 37 | 25 | 62 | 1.80 |
|  | > 50 | 54 | 66 | 120 |  |
|  |  | 91 | 91 | 182 |  |

## Discussion

To the best of our knowledge the present study is first s to use case control design to study the effect of Gaming on scholastic performance. Paucity of data on the effect of Gaming on scholastic performance of medical students makes comparisons difficult between other studies. In the present study 70(76.9%) of the cases were below twenty years which is similar to study conducted by Singh S ^7^ where majority of the participants were less than 22 yrs. Also in the study conduct by Chih-Hao Ku majority of participants were young adults^9^. In the present study Smartphone was the most commonly used gaming platform used by the study participants. Low cost of smartphones, better internet connectivity and ease of use can be a reason for smartphone preference in the present study.

In the present study there was no significant difference observed in the internal assessment scores and number of gaming hours. (p>0.05) i.e High gaming hours didn’t led to decrease in internal assessment score which is dissimilar to study done by Barry Ip et al where frequent gamers (both males and females) scored less than less frequent gamers in examinations with the average marks of non-gamers 9.4% higher than frequent gamers ^10^.

In the present study the internal assessment scores of Cases were 4.6% and 4.2% less than controls in 1^st^ and 2^nd^ internal assessments respectively. In the present study Male controls had a 9.7% & 7.4% more score than Male cases. This finding is somewhat similar to finding of Barry Ip et al where, examination marks of infrequent male gamers were averaged 7.2% higher than those for regular male gamers ^10^. Difference in scores of Female cases and controls were a meagre 0.4& 1.4 in 1s and 2^nd^ Internal assessment respectively. (p> 0.05).

This means, Gaming doesn’t affect scholastic performance in females. This finding is similar to study done by Barry Ip on gaming frequency and academic performance, where female students performed better than male students in all disciplines ^10^. In study conducted by Dumrique, D. O and associates, observed that academic performance of the school students was not affected even if they play online games ^11^.

In the present study Gaming addiction assessed by Lemmens Gaming Addiction Scoring (GAS) was found to be prevalent in 6.2% (using monothetic format) of study population (N) and 31% among cases (Nc). The prevalence of Gaming Disorder using different scales in various studies ranged from 2.0% to 22.7% ^8,12-24^. In specific studies, conducted by Lemmens et al the Gaming addiction was found to be 2.3% using monothetic format and 9.3% using Polythetic format of GAS ^8^. Also in study conducted by Mentzoni RA using GAS the prevalence of problematic users (≥4 of 7 on Game Addiction Scale) was 4.1% ^25^. In study conducted by Chong-Wen Wang in hongkong, 15.6% of study participants were identified as having a gaming addiction ^22^. In study conducted in Germany by Ruth Festl,3.7% of the respondents were considered as problematic Game users ^26^.

In the present study, there was a negative correlation observed between GAS and Mean internal assessment scores meaning that more the GAS scores less the internal assessment scores stressing the fact that gaming negatively affects Scholastic performance. In the review conducted by Mihara S, many studies reported lower educational, lower school grades and career attainment, in students indulging in gaming ^27^.

To the best of the of our knowledge this the first study to quantify the effect of Gaming on scholastic performance using a Case Control design. The present study found out that that the risk of low scores was (1.80-1.89) times more in cases than controls.

Limitations of the study: The study comes with the inherent limitations of the Case control design. The Findings of the present study pertains to a single educational setting which could limit its generalizability.

## Conclusion

The study concludes that Gaming adversely affects scholastic performance in males students but not in females. Awareness need to be created among medical students about the ill effects of Gaming which can have detrimental effect on their scholastic performance. Parents, teachers and peers should be made aware to detect Gaming addiction early. De -addiction services should be made available to those found having Gaming addiction in medical institutions. The study also opens new avenues for further exploration in different educational settings using Cohort study design.

Conflict of Interest: Deodatt M S, Divya R, Anju J, Raghuram V and Goyal R declare they have no competing interest

## Data Availability

Data is available on request to the author. Persons can visit the link below and access data on prior permission.

https://drive.google.com/file/d/11vF-rqCAGToutbGLSpG0rzb9e0K7AECP/view?usp=sharing

## Notes

### Competing Interest Statement

The authors have declared no competing interest.

### Author Declarations

Ethical principles such as respect for the person, and confidentiality were strictly adhered. Institutional Ethics clearance was obtained from Trichy SRM Medical College (IEC Code No: 1007/TSRMMCH&RC/ME-1/2019-IEC no :039)). Informed written consent was obtained from all the participants

